# Symptoms that predict positive COVID-19 testing and hospitalization: an analysis of 9,000 patients

**DOI:** 10.1101/2021.08.09.21261729

**Authors:** Mehrsa Jalalizadeh, Patricia A. F. Leme, Keini Buosi, Franciele A. V. Dionato, Cristiane F. Giacomelli, Luciana S.B. Dal Col, Karen L. Ferrari, Ana Carolina Pagliarone, Lucas M. Gon, Douglas F.O. Cezar, Akbar A. Esfahani, Leonardo O. Reis

**Affiliations:** UroScience, School of Medical Sciences, University of Campinas, UNICAMP and Pontifical Catholic University of Campinas, PUC-Campinas, Brazil

**Keywords:** symptoms, COVID-19, SARS-COV-2, diagnosis, hospitalization, prediction

## Abstract

**Purpose:** To develop a reliable tool that predicts which patients are most likely to be COVID-19 positive and which ones have an increased risk of hospitalization.

**Methods:** From February 2020 to April 2021, trained nurses recorded age, gender, and symptoms in an outpatient COVID-19 testing center. All positive patients were followed up by phone for 14 days or until symptom-free. We calculated the symptoms odds ratio for positive results and hospitalization and proposed a “random forest” machine-learning model to predict positive testing.

**Results:** A total of 8,998 patients over 16 years old underwent COVID-19 RT-PCR, with 1,914 (21.3%) positives. Fifty patients needed hospitalization (2.6% of positives), and three died (0.15%). Most common symptoms were: cough, headache, sore throat, coryza, fever, myalgia (57%, 51%, 44%, 36%, 35%, 27%, respectively). Cough, fever, and myalgia predicted positive COVID-19 test, while others behaved as protective factors. The best predictors of positivity were fever plus anosmia/ageusia (OR=6.31), and cough plus anosmia/ageusia (OR=5.82), both p<0.0001. Our random forest model had an ROC-AUC of 0.72 (specificity=0.70, sensitivity=0.61, PPV=0.38, NPV=0.86). Having steady fever during the first days of infection and persistent dyspnea increased the risk of hospitalization (OR=6.66, p<0.0001 and OR=3.13, p=0.003, respectively), while anosmia-ageusia (OR=0.36, p=0.009) and coryza (OR=0.31, p=0.014) were protective.

**Conclusion:** Present study and algorithm may help identify patients at higher risk of having SARS-COV-2 (online calculator http://wdchealth.covid-map.com/shiny/calculator/), and also disease severity and hospitalization based on symptoms presence, pattern, and duration, which can help physicians and health care providers.

## INTRODUCTION

The coronavirus disease 2019 (COVID-19) presents a variety of symptoms. Fever, cough, and myalgia are the most commonly reported symptoms since the description of the first cases from Wuhan [1]. In the following months, some articles discussed the epidemiology of the disease, its clinical features, and included reports about respiratory and non-respiratory symptoms such as gastrointestinal presentations, anosmia, and ageusia. Then, Kaye et al. presented the first report about anosmia, using the anosmia reporting tool for clinicians and found it to be present in 73% of patients, followed by cough (41%), fever (38%), headache (37%), and chills (27%) [2]. Those rates agree with many other reports [3]; however, most data come from in-hospital patients or office appointments. Considering that more than 80% of the patients develop mild disease [4] it is crucial to understand the symptoms in outpatients.

In 2020 Khan et al. [5] reported data from a COVID-19 screening clinic in Pakistan, including both inpatients and outpatients. They found that fever, cough, and shortness of breath were the most common presentations (72%, 59%, 57%, respectively). Although outpatients are included in many reports, a study focused on the outpatient population may bring light to the majority of cases at population. It could bring more reliable information about the symptom’s prevalence and relation to test positivity. In addition, it could present possible correlations with disease severity. That information could help clinicians and health care providers create protocols for COVID-19 testing and monitor high-risk patients.

This study aims to analyze data from all cases tested for COVID-19 using RT-PCR between February 2020 and April 2021 at the University of Campinas outpatient health center, Campinas, São Paulo, Brazil. The main objective is to develop a reliable tool that predicts which patients are most likely to be COVID-19 positive and which ones have an increased risk of hospitalization.

## METHODS

After local ethics committee approval number 4.173.069, all information was gathered from the University of Campinas outpatient health center, Campinas, São Paulo, one of the biggest universities in Brazil. All patients over 16 years old referred for COVID-19 RT-PCR testing were added to the database. Trained nurses recorded each patient’s data: age, gender, and symptoms. All positive patients were followed up by phone for 14 days or until symptom-free.

We calculated each symptom and symptoms combinations odds ratio for the probability of positive test results and uploaded the data of the symptoms into a “random forest” machine learning model to develop an algorithm that calculates their relation to positive COVID-19 results. We chose random forest over linear modeling due to the underlying nonlinear nature of symptoms, and also tested a linear model created by Menni et al. to test the reproducibility of their model in our data [6].

All statistical analyses were performed using R version 4.0.2 (2020-06-22) on RStudio platform version 1.3.1073 and using the following packages: tidyverse, lubridate, forecast, quantreg, splines, ggmap, pracma, and janitor. Normality for samples of n<5,000 was calculated using the Shapiro-Wilk test, and for n>5,000 the shape of the histogram was considered. Welch’s two-sample t-test was performed for parametric analysis of normal samples, and Fisher’s exact test for categorical data due to the small number of observations in some groups. An alpha error of less than 0.05 was considered a significance threshold for single testing. For multiple hypothesis testing in tables, we reduced the acceptable alpha (a) per hypothesis based on the number of tests (n) to keep the overall alpha error less than 10%: 0.1 = 1 - (1-a)^n [7]. The “Random Forest” model was created after dividing the data into train and test in 0.8:0.2 ratio, 100 models were trained and fitted. The best model was tested on the test set and receiver operating characteristic (ROC) was plotted. We used 10-fold cross-validation to avoid “overfitting” the model.

## RESULTS

From February 2020 to April 2021, a total of 8,998 COVID-19 RT-PCR tests were performed; a total of 1,914 (21.3%) of them returned positive, 70.6% returned negative, and 8% were either inconclusive or not available (NA). Six percent (n=117) of positive patients were asymptomatic and referred for testing due to exposure or screening. Patients with inconclusive or NA results were excluded for further analysis.

There was a gender discrepancy, with 70% of the patients being female. The mean age was 38 years old (minimum 16 and maximum 95).

### Positive PCR results

Patients with positive results were on average 1.8 years older (95%CI: 1.2 - 2.4 years, p<0.0001). The PCR samples were mostly collected 3 days after the start of the symptoms. There was no significant mean difference in the number of days between the start of symptom and sample collection between positive and negative patients (p = 0.23).

Table 1 describes the frequency of symptoms on the day of testing and the odds ratio for COVID-19 positivity. Interestingly, 72% of asymptomatic patients had positive PCR results. Only 27% of patients with cough, 22% of patients with headache, and 19% of patients with sore throats tested positive. The most common symptoms among the positive patients were cough (57%), headache (51%), sore throat (44%), coryza (36%), fever (35%), myalgia (27%), anosmia (11%), fatigue (8%), ageusia (7%), diarrhea (7%), and chills (2%). Rare symptoms included shortness of breath (1%), abdominal pain (<1%), loss of appetite (<1%), and nausea (<0.1%).

Considering isolated symptoms, anosmia and/or ageusia were the highest indicators of a positive result with the odds ratio of 5.46 (95%CI 4.43 – 6.7), followed by fever (1.93, 95%CI 1.72 – 2.16), cough (1.60, 95%CI 1.44 – 1.77), and myalgia (1.47, 95%CI 1.30 – 1.65), all with p<0.0001. On the other hand, presence of sore throat (OR=0.64, 95%CI 0.57 – 0.71 p<0.0001), headache (OR=0.88, 95%CI 0.80 – 0.98 p= 0.020), and coryza (OR=0.86, 95%CI 0.77 – 0.96, p=0.006) lowered the probability of positive result. However, due to multiple hypothesis testing, we suggest considering p values less than 0.004 as statistically significant and interpreting other p-values with caution.

**Table 1.** Symptom presentation based on test results. Due to multiple hypothesis testing we suggest considering p values less than 0.004 as significant (highlighted) and interpreting other p values with caution.

|  | **Positive**<br>**PCR**<br>**N=1,914** | **Negative**<br>**PCR**<br>**N= 6,358** | **OR** | **p-Value** |
| --- | --- | --- | --- | --- |
| **Female gender** | 1,301 (68%) | 4,479 (71%) | 1.12 | 0.041 |
| **Cough** | 1,086 (57%) | 2,867 (45%) | 1.60 | <0.0001 |
| **Sore throat** | 844 (44%) | 3,518 (55%) | 0.64 | <0.0001 |
| **Headache** | 971 (51%) | 3,419 (54%) | 0.88 | 0.020 |
| **Coryza** | 680 (36%) | 2,477 (39%) | 0.86 | 0.0067 |
| **Fever** | 666 (35%) | 1,377 (22%) | 1.93 | <0.0001 |
| **Myalgia** | 523 (27%) | 1,298 (20%) | 1.47 | <0.0001 |
| **Anosmia/Ageusia** | 247 (13%) | 168 (3%) | 5.46 | <0.0001 |
| **Dyspnea** | 14 (<1%) | 66 (1%) | 0.70 | 0.29 |
| **Chills** | 42 (2%) | 121 (2%) | 1.16 | 0.45 |
| **Fatigue** | 158 (8%) | 622 (10%) | 0.83 | 0.045 |
| **Diarrhea** | 136 (7%) | 887 (14%) | 0.47 | <0.0001 |
| **Abdominal pain** | 6 (<1%) | 50 (<1%) | - | - |
| **Loss of appetite** | 14 (<1%) | 32 (<1%) | - | - |
| **Nausea** | 3 (<1%) | 86 (<1%) | - | - |
| **Vomiting** | 0 | 3 (<1%) | - | - |
| **Fever + Cough** | 460 (24%) | 640 (10%) | 2.83 | <0.0001 |
| **Fever + Cough + Dyspnea** | 2 (<1%) | 3 (<1%) | - | - |
| **Fever + Anosmia/Ageusia** | 70 (4%) | 38 (<1%) | 6.31 | <0.0001 |
| **Fever + Chills** | 12 (<1%) | 35 (<1%) | - | - |
| **Fatigue + Myalgia** | 189 (10%) | 319 (5%) | 2.07 | <0.0001 |
| **Cough +**<br>**Anosmia/Ageusia** | 126 (7%) | 76 (<1%) | 5.82 | <0.0001 |
| **Headache + Coryza** | 386 (20%) | 1,303 (20%) | 0.98 | 0.78 |
| **Cough + Sore throat** | 589 (31%) | 1,794 (28%) | 1.13 | 0.033 |
| **Asymptomatic** | 117 (6%) | 25 (<1%) | 16.50 | <0.0001 |

The majority of patients complained of 3 symptoms, and **Figure 1a** shows the distribution of the count of symptoms whereas **Figure 1b** shows the proportion of positive testing based on how many concurrent symptoms each patient has; there is a slightly increased proportion of positive cases with an increased number of concurrent symptoms although patients with 6 or more concurrent symptoms were very rare in our data. **Figure 2** shows symptoms distributions by age. The age plot indicates that older patients would consider getting tested with fewer symptoms compared to younger patients who needed more presenting symptoms to consider COVID-19 testing.

**Figure 1.**
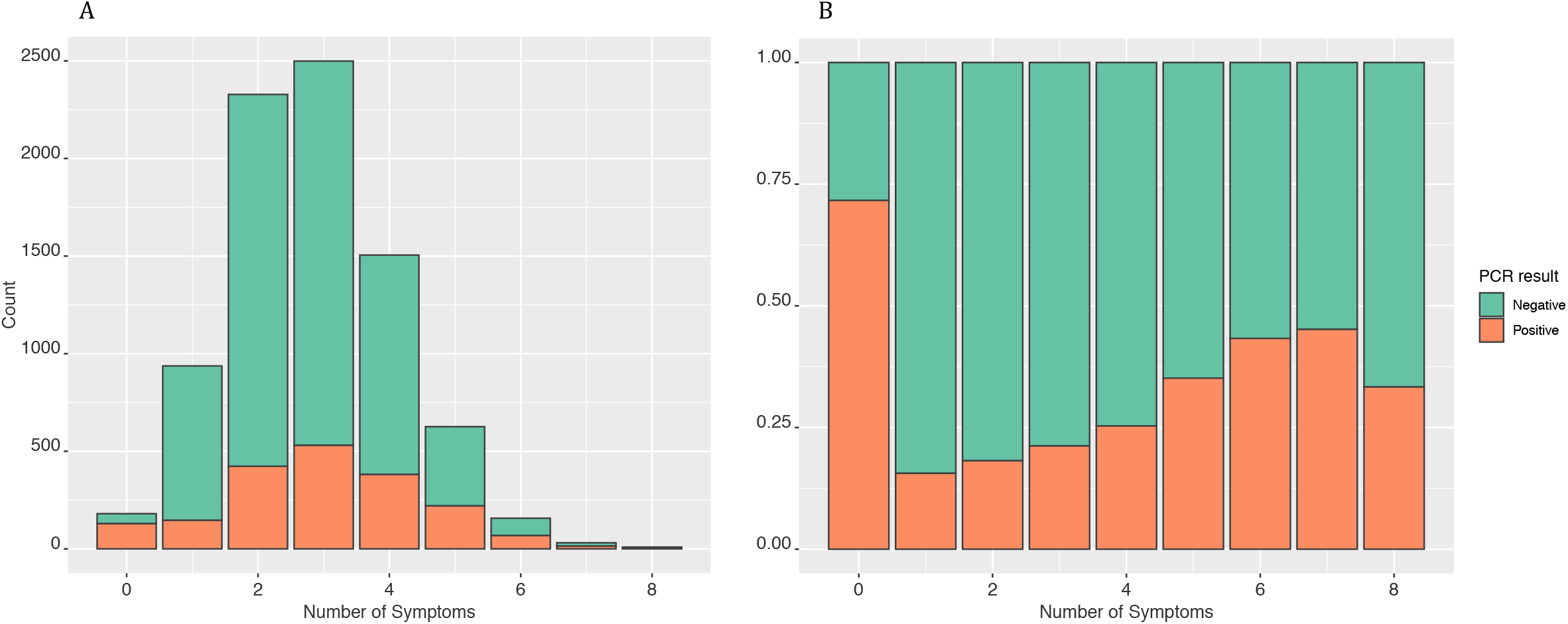
**a.** Count of tested individuals based on the number of concomitant symptoms. **b**. Percent positive results based on the number of concomitant symptoms. Individuals with zero symptoms (asymptomatic) were referred due to exposure or screening. Interesting, 72% of them were positive.

**Figure 2.**
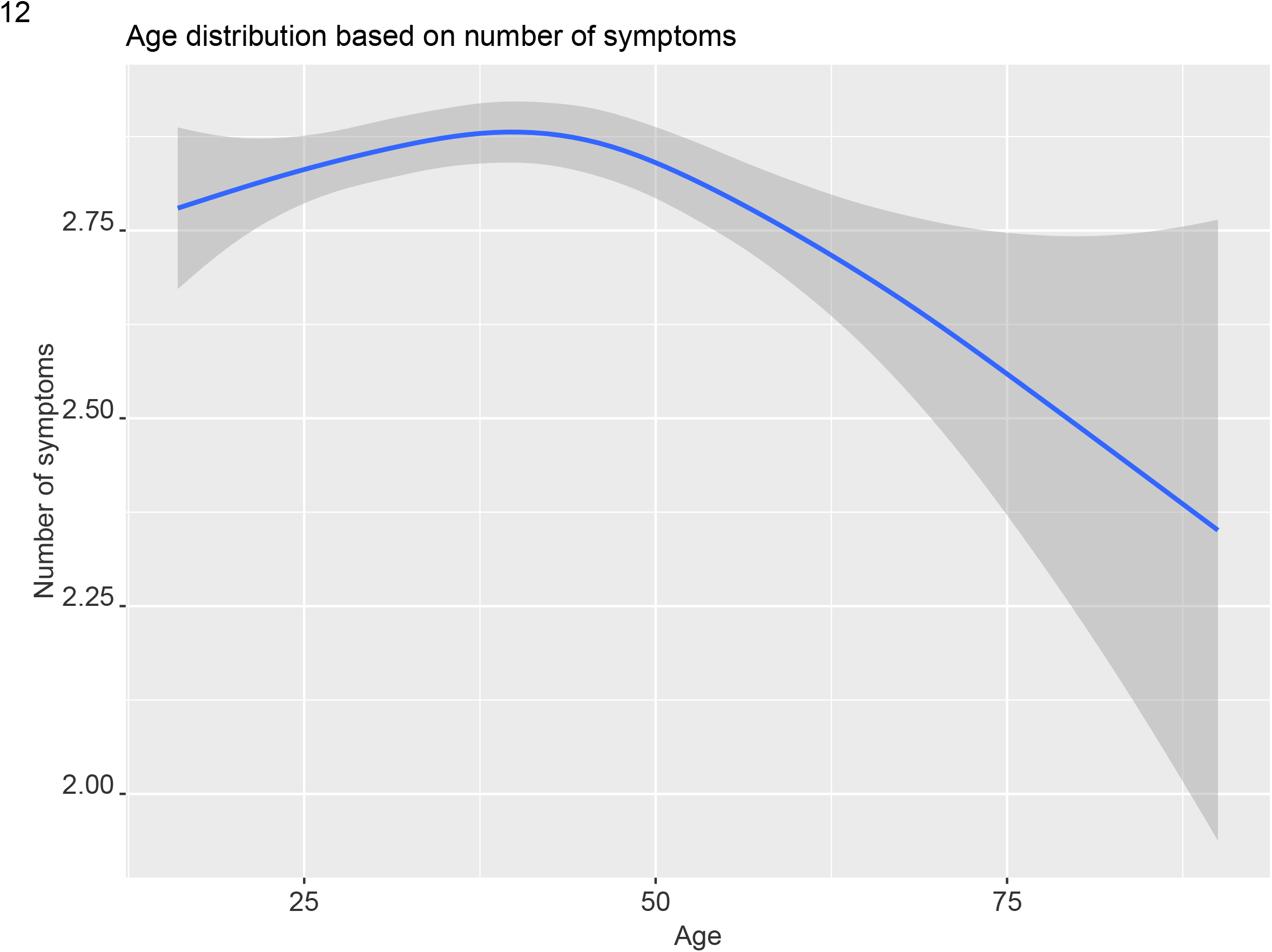
Age distribution based on the number of concomitant symptoms. Younger individuals are referred for testing with more concomitant symptoms.

The correlation plot in **Figure 3** shows the likelihood of each two symptoms co-presenting in a patient. Anosmia had a high correlation with ageusia (73%, p<0.005); therefore, the two symptoms were considered dependent and were used concomitantly in further analyses. The presence of anosmia and ageusia was slightly correlated with the concomitant presence of coryza and headache.

**Figure 3.**
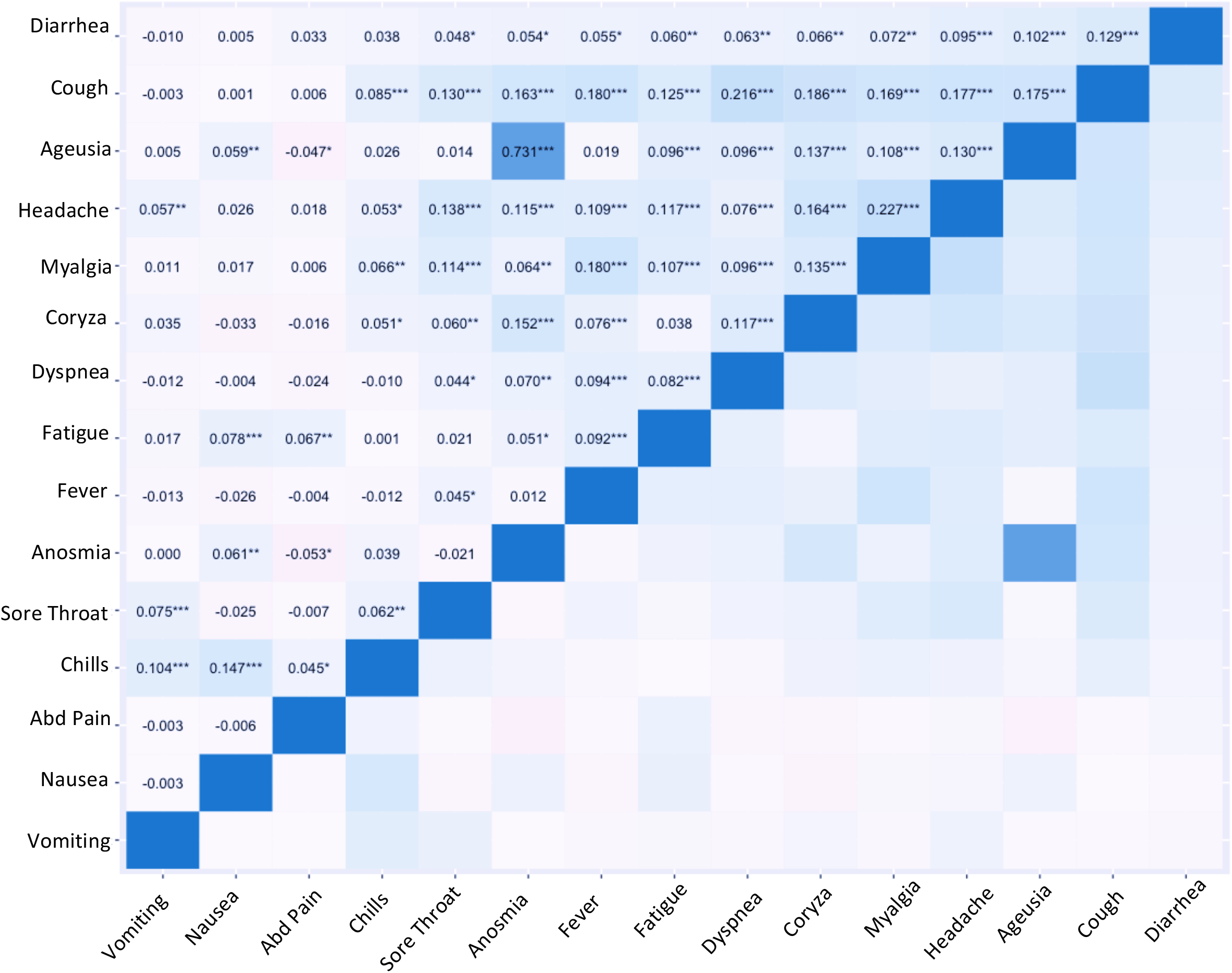
Correlation plot showing the likelihood of each two symptoms co-representing in each patient. * p-value between 0.05 and 0.005; ** p-value between 0.005 and 0.0005; *** p-value less than 0.0005.

Considering symptoms combination, the highest odds ratio came from fever plus anosmia/ageusia (OR=6.31, 95%CI 4.24 – 9.40), followed by cough plus anosmia/ageusia (5.82, 95%CI 4.36 – 7.78), and fever plus cough (2.83, 95%CI 2.47 – 3.23), all p<0.0001 (**Table 1**). The former combination of symptoms, although having a high odds ratio of positivity, was very rare: only 4% of positive patients.

**Figures 4** and **5** show our random forest model’s ROC curve and the most important symptoms predictive of positive testing. The final model used 2000 trees (mtry=2, min_n=29) on 10-fold cross validation. Area under the curve of our model (ROC-AUC) was 0.72 with an accuracy of 0.68 (sensitivity = 0.61, specificity = 0.70, positive predictive value [PPV] = 0.38, negative predictive value [NPV] = 0.86). We developed an online calculator based on our model’s algorithm, which is currently available at http://wdchealth.covid-map.com/shiny/calculator/. We tested linear model created by Menni et al. [6] on our data which revealed an ROC-AUC of 0.602 (**Figure 6**), with sensitivity = 0.23, specificity = 0.93, PPV = 0.50, and NPV = 0.80.

**Figure 4.**
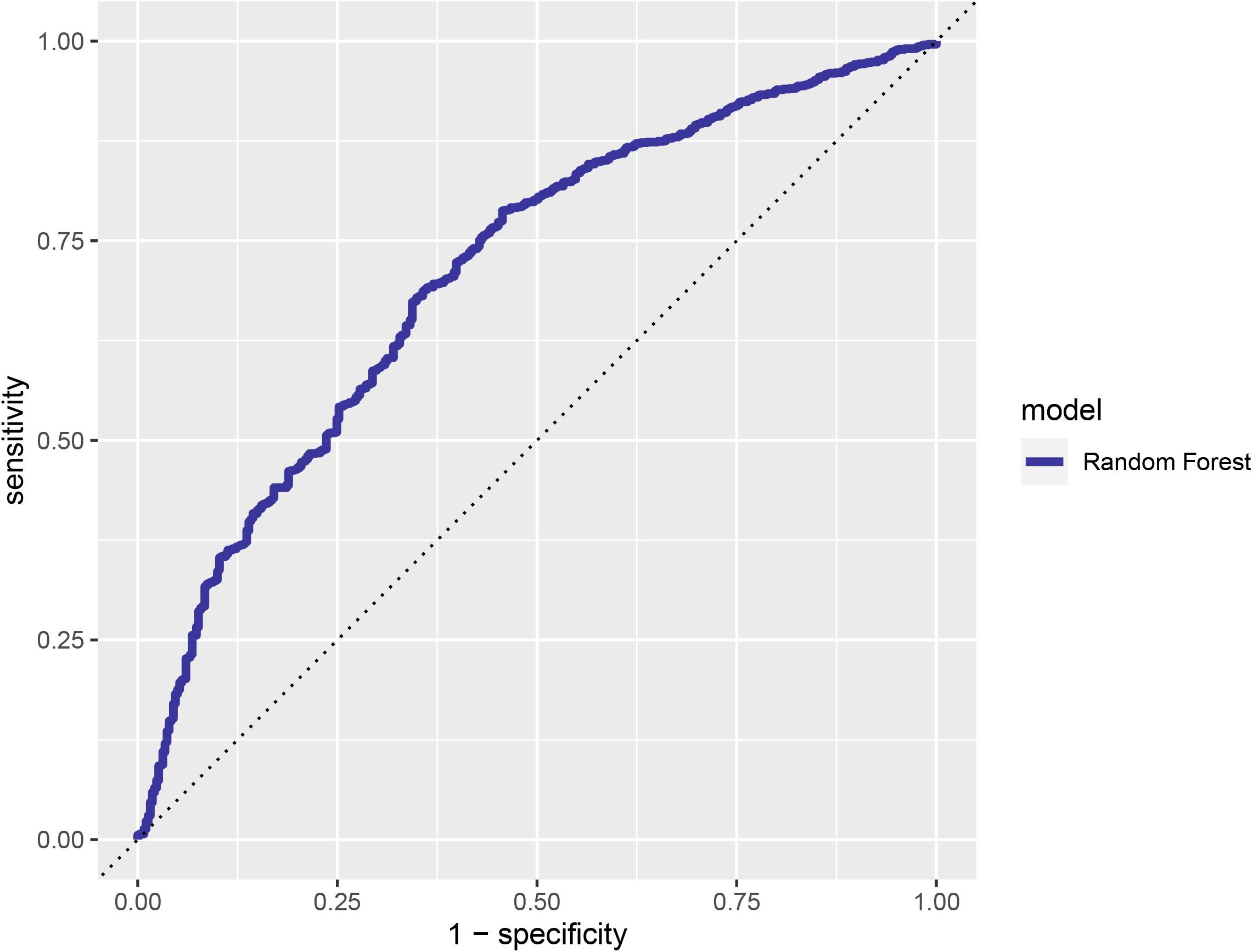
ROC of our random forest model with AUC of 0.72, and accuracy of 0.68 (sensitivity = 0.61, specificity = 0.70, PPV = 0.38, NPV = 0.86).

**Figure 5.**
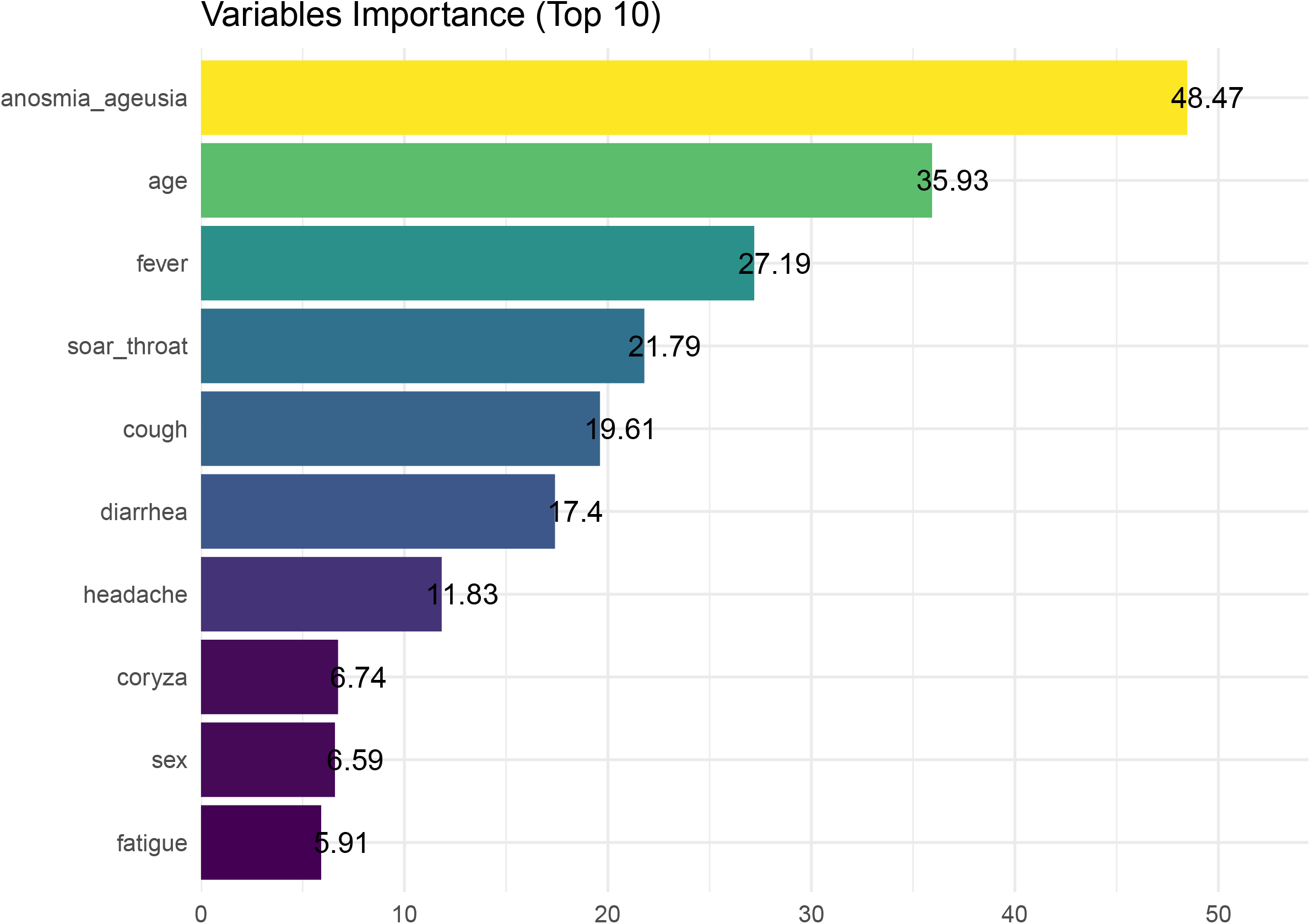
Importance of variables in our random forest model.

**Figure 6.**
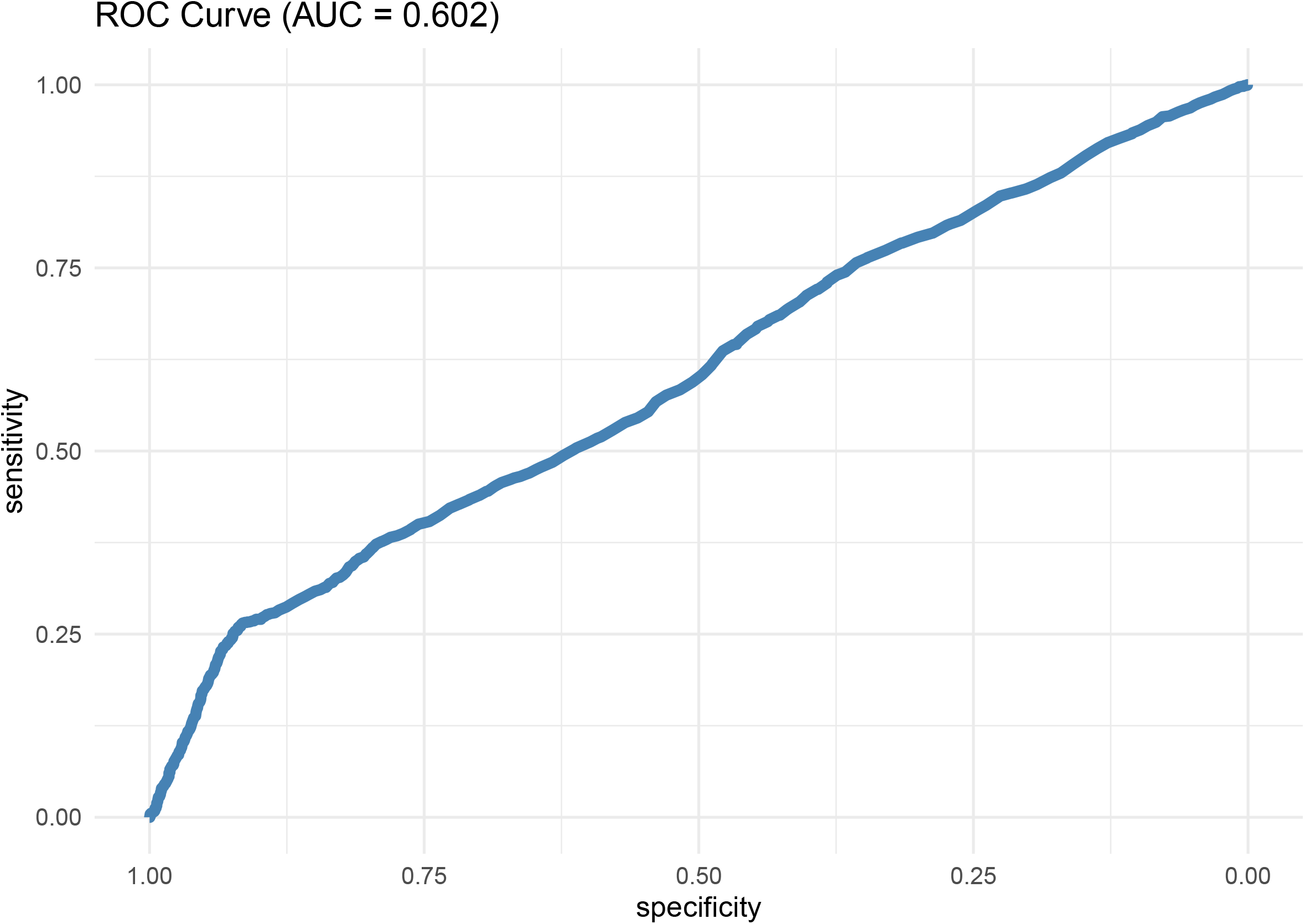
ROC of Menni et al linear regression model on our data shows very low accuracy AUC = 0.602 sensitivity = 0.23, specificity = 0.93, PPV = 0.50, and NPV = 0.80.

### Risk of hospitalization

Fifty patients needed hospitalization (2.6% of all positive cases), and three died (0.15%). **Table 2** describes overall symptoms presentation among COVID-19-positive patients: starting symptoms and emerging new symptoms throughout the disease and their association with hospitalization.

**Table 2.** Symptom presentation based on hospitalization. Due to multiple testing, we suggest considering p values less than 0.006 as significant (highlighted) and interpreting other p values with caution.

|  | **Hospitalized**<br>**N=50** | **Outpatient**<br>**N= 2045** | **OR** | **p-Value** |
| --- | --- | --- | --- | --- |
| **Fever (first 5 days)** | 20 (40%) | 186 (9%) | **6.66** | **<0.0001** |
| **Fever (days 6 to 11)** | 5 (10%) | 170 (8%) | 1.22 | 0.67 |
| **Dyspnea (first 5 days)** | 9 (18%) | 184 (9%) | 2.22 | 0.0340 |
| **Dyspnea (days 6 to 11)** | 9 (18%) | 134 (7%) | 3.13 | **0.0026** |
| **Chills** | 3 (6%) | 26 (1%) | 4.96 | 0.0107 |
| **Coryza** | 5 (10%) | 537 (26%) | 0.31 | 0.0140 |
| **Sore throat** | 7 (14%) | 418 (20%) | 0.63 | 0.27 |
| **Cough** | 25 (50%) | 764 (37%) | 1.67 | 0.071 |
| **Headache** | 15 (30%) | 632 (31%) | 0.96 | 0.89 |
| **Fatigue** | 15 (30%) | 337 (16%) | 2.17 | 0.013 |
| **Anosmia/Ageusia** | 8 (16%) | 706 (35%) | 0.36 | 0.0088 |
| **Diarrhea** | 3 (6%) | 126 (6%) | 0.97 | 0.96 |
| **Loss of appetite** | 3 (6%) | 47 (2%) | 2.90 | 0.10 |
| **Abdominal pain** | 1 (2%) | 11 (<1%) | 3.76 | 0.25 |
| **Nausea** | 1 (2%) | 12 (<1%) | 3.45 | 0.27 |
| **Vomiting** | 0 | 3 (<1%) | - | - |
| **Myalgia** | 17 (34%) | 401 (21%) | 2.11 | 0.0138 |
| **Asymptomatic on day 1** | 3 (6%) | 173 (8%) | 0.69 | 0.54 |

Patients with steady fever during the first five days were significantly more likely to get hospitalized (OR = 6.66, 95%CI 3.71 – 11.97, p < 0.0001). Indeed, 40% of patients who needed hospitalization had a fever in the first five days of the disease, while only 9% of outpatients had it. The presence of dyspnea during the first five days also increased the risk of hospitalization (OR = 2.22, 95%CI 1.06 – 4.64, p = 0.0340). If this symptom was persistent for 11 days, it increased the risk of hospitalization even further (18% of the hospitalized vs. 7% of outpatients, OR = 3.13, 95%CI 1.49 – 6.58, p=0.0026).

In contrast, patients who developed fever on days six to eleven had no statistically significant risk of hospitalization (OR = 1.22, 95%CI 0.48 – 3.13, p=0.67). Interestingly, patients with anosmia/ageusia or coryza had lower odds of hospitalization (anosmia-ageusia: OR=0.36, 95%CI 0.17 – 0.77, p = 0.009, coryza: OR = 0.31, 95%CI 0.12 – 0.79, p = 0.014).

## DISCUSSION

This is one of the largest cohorts of RT-PCR COVID-19 tests in the general population to date. Most of the currently published literature regards hospitalized patients even though the majority of cases are mild. Therefore, there is a lack of information about symptoms prevalence and relation to the severity in general cases. Indeed, about 80% of COVID-19 patients have mild symptoms. Our data present important findings in the majority of COVID-19 patients who do not need medical assistance or hospitalization, including symptoms frequencies and their potential in predicting positivity or severity by hospitalization need.

The most common symptoms in our data were: cough, headache, sore throat, coryza, fever, myalgia, (57%, 51%, 44%, 36%, 35%, and 27%, respectively) which is close to previously published studies [1-5]. Cough, fever, and myalgia were more common in the positive COVID-19 patients, while sore throat, headache, and coryza behave as protective factors indicating that the latter symptoms are more likely due to non-COVID-19 causes (allergies, common cold, etc.). Patients with two or more concurrent symptoms were slightly more likely to test positive for COVID-19.

Anosmia or ageusia was present in only 13% of positive cases at the time of testing. However, their presence was significantly associated with positive test results. It increased the risk of positive testing by 5.5-fold, while fever increased it by 1.9 and cough only 1.6 times. Of note, patients with a sore throat, diarrhea, and coryza were less likely to test positive, indicating that other causes are more likely in these symptoms (other viral illnesses, allergies, food poisoning, etc.).

There is not one single sign or symptom that can predict the positivity for COVID-19. Menni et al. [6] proposed an algorithm that includes gender, age, anosmia, cough, severe fatigue, and skipped meals. We tested the algorithm on our data, and it revealed a ROC-AUC of 0.6 with a very low sensitivity of 0.23. Their model relies heavily on anosmia and ageusia, which were rare symptoms (only 13%); this explains its low sensitivity. In addition, their model is based on self-reported symptoms, while, in our study, trained nurse practitioners collected data. Our data present more negative than positive results, and this imbalance may explain the lower AUC when running Menni et al. model. However, our model is a better reflection of the general population compared to the Menni et al. model, which will always overestimate the rate of positive results. Moreover, the oversimplification of the approach (using a linear model) might generate hysteria in the population, causing further distrust of the scientific community. We know that the relationships between symptoms and COVID-19 diagnosis and severity are not linear and the work of Menni et al. is a great example of oversimplifying a complex problem.

We used “random forest’ modeling with 2000 trees and 10-fold cross-validation that was trained on 80% of the data and finally tested on the remaining 20% of the data. We avoided “over-fitting” the model since our data is not a true representative of all populations (limited geographic location, high female ratio, etc.). The area under the curve of our model was 0.72 with an accuracy of 0.68 (sensitivity = 0.61, specificity = 0.70, PPV = 0.38, NPV = 0.86). Our online calculator at http://wdchealth.covid-map.com/shiny/calculator/ can inform single individuals of their risk and the features contributing to it. It could also guide decision-making and discussions with the healthcare providers.

Fever and dyspnea had predictive potential in calculating the disease severity and risk of hospitalization. Interestingly, the pattern and duration of these symptoms had major significance rather than their presence. Patients who presented a steady fever early in the disease had a significantly higher chance of hospitalization. Fever is a sign of more severe illness and indicates a systemic inflammatory response. In the early stages of the disease, it may show an inefficient virus control, which can cause tissue damage by the inappropriately high response to the presence of the virus, eventually leading to organ failure. According to previous studies, fever, malaise, and dry cough are frequent in the initial stage of COVID-19, when there is invasion and infection of the upper respiratory tract, with a greater immune response involving the release of C-X-C motif chemokine ligand 10 (CXCL-10) and interferons (IFN-β and IFN-λ) from the virus-infected cells. The majority of patients do not progress beyond this phase as the mounted immune response is sufficient to contain the spread of infection [8].

However, steady fever in the early stages may be a sign of virus invasion at type 2 alveolar epithelial cells, releasing a systemic inflammatory response that is inefficient in controlling the virus and may cause subsequent inflammation and lung injury. On the other hand, the late onset of fever in COVID-19 disease may indicate the development of a secondary bacterial infection (e.g., sinusitis). Therefore, instead of a progression of coronavirus to the lower respiratory tract, it may indicate bacterial contamination of the upper respiratory tract, which does not mean a worse prognosis. Bacterial respiratory tract infection can be efficiently treated with antibiotics in the majority of patients.

Patients with a long duration of dyspnea also had a higher chance of hospitalization. This can be explained either by respiratory fatigue in these patients or long-term persistence of infection and inflammation in their lungs leading to accumulative tissue damage as time passes. Mechanisms of hypoxemia in COVID-19 are partially described by Dhont et al. [9]. Arterial hypoxemia early in SARS-CoV-2 infection is primarily caused by V/Q mismatch, resulting in intrapulmonary shunting. As the disease progresses, further loss of lung perfusion regulation, intravascular microthrombi, and impaired lung diffusion capacity occurs, and dyspnea becomes more apparent, leading to increased risk of hospitalization.

Our finding that anosmia/ageusia and coryza decrease the likelihood of hospitalization could indicate that the virus probably stays in the nasal area in these patients. Of note, the presence of anosmia and ageusia was slightly correlated with the concomitant presence of coryza and headache. The nasal congestion by itself may decrease the sense of smell, independently from COVID-19’s direct effect on the olfactory nerves. Moreover, the virus in the upper respiratory region causes a more manageable disease compared to lung disease.

Our study reflects a population of a big city in Brazil and it may not reflect other places. The study’s population had a high proportion of females, young people, and healthcare workers (further explored in another article). However, it brings evidence about the general population, which in the high majority does not require hospitalization, and thus literature has limited information about it. The online calculator will be updated according to population previous infection, virus variants and vaccination impact.

## CONCLUSION

The present study and algorithm may help identify patients at higher risk of having SARS-COV-2 (online calculator http://wdchealth.covid-map.com/shiny/calculator/), and also disease severity and hospitalization based on symptoms presence, pattern, and duration, which can help physicians and health care providers.

## Data Availability

The data that support the findings of this study are available from the corresponding author upon reasonable request.

http://wdchealth.covid-map.com/shiny/calculator/

## Financial Disclosure

The authors declare that they have no relevant financial interests.

## Author Contributions

MJ, LMG, and AAE: data analysis and manuscript writing.

PAFL, KB, FAVD, CFG, LSBDC, KLF, ACP, DFOC: data collection and manuscript editing LOR: funding acquisition, project development, supervision.

## Funding Support

Coordination for the Improvement of Higher Education Personnel-CAPES: 88887.506617/2020-00 and National Council for Scientific and Technological Development – CNPq, Research Productivity: 304747/2018-1

The funder had no involvement in study design, data collection, data analysis, manuscript preparation, and/or publication decisions.

## Acknowledgment

To the involved institution(s), the patients, and those that provided and cared for study patients.

## Conflict of Interests

None declared.

## Research Involving Human Participants

University of Campinas ethics committee approval number: 4.173.069

## Notes

### Competing Interest Statement

The authors have declared no competing interest.

### Author Declarations

University of Campinas ethics committee approval number: 4.173.069

## References

1- Huang C, Wang Y, Li X, et al. Clinical features of patients infected with 2019 novel coronavirus in Wuhan, China [published correction appears in Lancet. 2020 Jan 30]. Lancet. 2020;395(10223):497–506. doi:10.1016/S0140-6736(20)30183-5.

2- Kaye R, Chang CWD, Kazahaya K, Brereton J, Denneny JC 3rd. COVID-19 Anosmia Reporting Tool: Initial Findings. Otolaryngol Head Neck Surg. 2020;163(1):132–134. doi:10.1177/0194599820922992.

3- Esakandari, H., Nabi-Afjadi, M., Fakkari-Afjadi, J., Farahmandian, N., Miresmaeili, S. M., & Bahreini, E. (2020). A comprehensive review of COVID-19 characteristics. Biological procedures online, 22, 19. https://doi.org/10.1186/s12575-020-00128-2.

4- Ortiz-Prado, E., Simbaña-Rivera, K., Gómez-Barreno, L., Rubio-Neira, M., Guaman, L. P., Kyriakidis, N. C., Muslin, C., Jaramillo, A., Barba-Ostria, C., Cevallos-Robalino, D., Sanches-SanMiguel, H., Unigarro, L., Zalakeviciute, R., Gadian, N., & López-Cortés, A. (2020). Clinical, molecular, and epidemiological characterization of the SARS-CoV-2 virus and the Coronavirus Disease 2019 (COVID-19), a comprehensive literature review. Diagnostic microbiology and infectious disease, 98(1), 115094. https://doi.org/10.1016/j.diagmicrobio.2020.115094.

5- Khan, M., Khan, H., Khan, S., & Nawaz, M. (2020). Epidemiological and clinical characteristics of coronavirus disease (COVID-19) cases at a screening clinic during the early outbreak period: a single-center study. Journal of medical microbiology, 69(8), 1114–1123. https://doi.org/10.1099/jmm.0.001231.

6- Menni C, Valdes AM, Freidin MB, et al. Real-time tracking of self-reported symptoms to predict potential COVID-19. Nat Med. 2020;26(7):1037–1040. doi:10.1038/s41591-020-0916-2.

7- Beran, J., 1994. Statistics for Long-Memory Process, Boca Raton: CRC Press LLC.

8- Parasher A. COVID-19: Current understanding of its Pathophysiology, Clinical Presentation, and Treatment. Postgrad Med J. 2021 May;97(1147):312–320.

9- Dhont S, Derom E, Van Braeckel E, Depuydt P, Lambrecht BN. The pathophysiology of ’happy’ hypoxemia in COVID-19. Respir Res. 2020 Jul 28;21(1):198. doi: 10.1186/s12931-020-01462-5. PMID: 32723327; PMCID: PMC7385717.

